# Prospective predictors of risk and resilience trajectories during the early stages of the COVID-19 pandemic: a longitudinal study

**DOI:** 10.1101/2021.10.08.21264752

**Authors:** Tal Shilton, Anthony D Mancini, Samantha Perlstein, Grace E Didomenico, Elina Visoki, David M Greenberg, Lily A Brown, Raquel E Gur, Rebecca Waller, Ran Barzilay

**Author notes:** Corresponding author: Ran Barzilay MD PhD, 10th floor, Gates Building, Hospital of the University of Pennsylvania, 34th and Spruce Street, Philadelphia, PA 19104. These authors contributed equally to manuscript preparation (Tal Shilton, Anthony D Mancini).

## Abstract

**Background:** The COVID-19 pandemic is a rapidly evolving stressor with significant mental health consequences. We aimed to delineate distinct anxiety-response trajectories during the early stages of the pandemic and to identify baseline risk and resilience factors as predictors of anxiety responses.

**Methods:** Using a crowdsourcing website, we enrolled 1,362 participants, primarily from the United States (n = 1064) and Israel (n = 222) over three time-points from April-September 2020. We used latent growth mixture modeling to identify anxiety trajectories over time. Group comparison and multivariate regression models were used to examine demographic and risk and resilience factors associated with class membership.

**Results:** A four-class model provided the best fit. The resilient trajectory (stable low anxiety) was the most common (n = 961, 75.08%), followed by chronic anxiety (n = 149, 11.64%), recovery (n = 96, 7.50%) and delayed anxiety (n = 74, 5.78%). While COVID-19 stressors did not differ between trajectories, resilient participants were more likely to be older, living with another person and to report higher income, more education, fewer COVID-19 worries, better sleep quality, and more dispositional resilience factors at baseline. Multivariate analyses suggested that baseline emotion regulation capabilities and low conflictual relationships uniquely distinguished participants in distinct trajectories.

**Conclusions:** Consistent with prior resilience research following major adversities, a majority of individuals showed stable low levels of low anxiety in response to the COVID-19 pandemic. Knowledge about dispositional resilience factors may prospectively inform mental health trajectories early in the course of ongoing adversity.

---

At the end of January 2021, a year into the COVID-19 pandemic, nearly 100 million cases of COVID-19 have been reported globally, including more than 2 million deaths (Wold Health Organization, 2021). The rapidly evolving situation contributed to compulsory measures that have drastically altered people’s lives worldwide, with substantial potential impacts on health and wellbeing (Cusinato et al., 2020; Evanoff et al., 2020). There has also been widespread attention paid to the evolving longitudinal burden on population-wide mental health (Gordon & Borja, 2020), highlighting the urgent need to improve our understanding of the variability in mental health responses during and following COVID-19.

Prior research suggests that the majority of people exhibit stable levels of functioning after exposure to various forms of acute adversity (Mancini, Bonanno, & Clark, 2011), including after outbreaks of high-risk infectious diseases (Bonanno et al., 2008; Bults et al., 2011; Leung et al., 2005). However, stress responses typically show substantial individual variation (Bonanno & Mancini, 2012), and thus it is critical to account for inter-individual variability in response to COVID-19 stress. The empirical literature suggests that this variability can be captured by a relatively small set of prototypical outcome trajectories (Bonanno 2004). The most common trajectory of resilience is characterized by a relatively stable trajectory of healthy functioning even soon after an acute stressor. By contrast, a recovery pattern is distinguished by elevated symptoms and a gradual return to normal levels of functioning, while chronic distress is characterized by elevated symptoms that persist. Finally, delayed distress is characterized by moderate to elevated symptoms that worsen over time. Given that the effect of COVID-19 on mental health will depend on a wide variety of individual and contextual factors, it is critical to use techniques that can identify these individually varying response patterns (Mancini, 2020). However, relatively few studies have examined individual variability in response to the pandemic’s early stress, when anxiety and uncertainty may have been most acute.

In addition to variation at the individual level, the pandemic itself has undergone rapid and unstable changes in virus spread leading to both imposition and relaxation of government restrictions, which together may have affected mental health trajectories since the predictability and capacity to control a stressor are critical to individual adjustment (Cheng, Lau, & Chan, 2014). Longitudinal studies using pre-pandemic data suggested negative effects on mental health symptoms during the initial stages of the COVID-19 pandemic (Shi et al. 2020; Xiong et al. 2020; Ma et al. 2020; Barzilay et al. 2020; Kwong et al. 2020; Pierce et al. 2020; Vindegaard and Eriksen Benros 2020) and a few longitudinal studies conducted at the early stages of the pandemic (March to May) demonstrated an increase in psychological distress, with elevated rates of anxiety, depression, and suicide risk (O’Connor et al., 2020; Ruggieri, Ingoglia, Bonfanti, & Lo Coco, 2020). Nevertheless, later and more recent reports suggested that anxiety symptoms subsequently declined or were small in magnitude (Daly, Sutin, & Robinson, 2020; McGinty, Presskreischer, Anderson, Han, & Barry, 2020; Prati & Mancini, 2021) and even recovered to pre-pandemic levels (Daly & Robinson, 2020; Robinson & Daly, 2020). Taken together, it remains unclear how mental health trajectories during the pandemic have evolved over time (Mancini, 2020). Thus, there remains a critical need for longitudinal studies of adjustment during the early stages of COVID-19 using data with sufficient baseline phenotyping to allow for identification of factors that contribute to both resilient and adverse mental health trajectories.

In the current study, we sought to address these knowledge gaps while overcoming traditional challenges in resilience research by integrating both trait and outcome approaches. Specifically, we assessed dispositional resilience factors at baseline and then assessed their capacity to account for individual variation in resilient and other trajectories of adjustment. We used data collected through a research crowdsourcing website (Barzilay et al., 2020) that allowed for (1) longitudinal modeling of mental health trajectories over time using repeated assessment of anxiety symptoms; (2) identification of a trajectory of healthy functioning and positive adaptation after exposure to adversity (Bonanno, 2004); and (3) examination of baseline risk and resilience factors that may have contributed to belonging to each trajectory. We included a multidimensional assessment of resilience factors using a battery that probed intrapersonal, interpersonal, and social environment characteristics (Moore et al., 2020), in accordance with widely accepted method encouraging a multi-level approach to evaluate both an individual’s traits and resources (Cicchetti & Curtis, 2007; Luthar, Cicchetti, & Becker, 2000). We explored the possibility that; (1) participants would belong to heterogeneous subpopulations that comprised distinct anxiety response trajectories over time, with a resilient trajectory being the most prevalent (Leung et al., 2005; Mancini et al., 2011); and (2) resilience and risk factors would be associated with individually-varying trajectories. We were particularly interested in identifying whether demographic factors, COVID-19 stressors and worries, and dispositional resilience factors predicted participant’s trajectories membership.

## Methods

### Participants and Procedures

On April 6th 2020, we launched a website (https://www.covid19resilience.org/) that included a resilience survey, assessment of COVID-19-related stress (worries) and mental health screening (Barzilay et al., 2020). At the end of the survey, participants received feedback on their resilience scores with personalized recommendations regarding stress management. The feedback functioned to incentivize participants to complete the survey carefully. Following the feedback, participants were asked if they are interested in being re-contacted for future surveys. The study was advertised through: 1) Researchers’ social networks, including emails to colleagues around the world; 2) Social media; 3) The University of Pennsylvania and Children’s Hospital of Philadelphia internal notifications and websites; and 4) Organizational mailing lists. All participants consented to participate in the study. The study was approved by the Institutional Review Board of the University of Pennsylvania.

Analyses included 1,392 participants above the age of 18 who had completed at least two out of three time points for the anxiety screening: T1, April 6th to May 10th, *n* = 1289; T2, May 12^th^ to July 6th, *n* = 1,200; and T3, August 25^th^ to - September 27^th^, *n* = 914. Overall, 448 participants had T1 and T2 data only, 162 had T1 and T3 only and 73 had T2 and T3 data only. Six hundred and seventy-nine participants, 49.8%, had complete data across all three time points and 50.2% had data available for at least 2 time-points.

### Measures

#### Anxiety symptoms

Participants completed the Generalized Anxiety Disorder 7 (GAD-7) questionnaire, a self-report scale developed to assess the defining symptoms of anxiety (Spitzer, Kroenke, Williams, & Löwe, 2006). The items are rated on a 4-point Likert-type scale (from 0□=□not at all to 3□=□nearly every day) and scores ranged from 0 to 21. In the current study, the Cronbach’s alpha coefficient of the GAD-7 was .89 at T1, .92 at T2 and .92 at T3.

#### COVID-19-related worries

Participants indicated the degree to which they were worried about a variety of COVID-19 related outcomes on 5-point Likert scale (0 = Not at all; 4 = a great deal). Worries included: 1) Contracting COVID-19; 2) Dying from COVID-19; 3) Family members contracting COVID-19; 4) Unknowingly infecting others with COVID-19; 5) Currently having COVID-19; 6) Having significant financial burden because of the COVID-19 pandemic.

#### COVID-19-related stressors

Participants were asked to rate whether they had experienced the following: 1) Testing positive for COVID-19; 2) knowing someone who died from COVID-19; 3) job loss since the start of the COVID-19 pandemic; and 4) reduced pay since the start of the COVID-19 pandemic. These variables were scored as Yes = 1 (experienced), No = 0 (not experienced). The stressors were summed for a Cumulative stressors measure.

#### Pre-existing anxiety diagnosis

Participants were asked whether they had received a diagnosis of generalized anxiety disorder prior to the COVID-19 pandemic (1 = yes, 0 = no).

#### Sleep measure

Participants completed the Insomnia Severity Index (ISI), a 7-item assessment of insomnia symptoms over the prior two weeks, with items rated on a scale ranging from 0 (No problems) to 4 (Very Severe) (Morin, Belleville, Bélanger, & Ivers, 2011).

#### Self-reported resilience factors

The survey included 21 items assessing resilience factors that were recently compiled into a single battery (Barzilay et al., 2020). The questions were selected from a larger set of 212 items using factor analysis followed by computerized adaptive test simulation (Moore et al., 2020). The survey included five subscales of resilience factors: self-reliance (3 items; e.g., can usually find a way out of difficult situations), with items coded on a 7-point Likert Scale (1 = strongly disagree to 7 = strongly agree); emotion regulation (5 items; difficulty concentrating or controlling behaviors when upset, limited access to emotion regulation strategies), with items coded on a 5-point Likert Scale (1 = almost always [91-100%] to 5 = almost never [0-10%]); characteristics of close relationships: positive (4 items on supportive close relationships; e.g., lasting relationship and level of care) and negative (5 items on hostility in relationships; e.g., level of arguing), with items coded on a 5-point Likert Scale (1 = little or none to 5 = the most);); and perceptions of the neighborhood environment (4 items; e.g., perceived level of trust and safety in neighborhood), with items coded on an 5-point Likert Scale (1 = strongly disagree to 5 = strongly agree). To maximize interpretability, we coded all items such that a higher score indicated greater resilience.

### Analytic Strategy

To address our first study aim, we applied latent growth mixture modeling (LGMM) to the anxiety scores at T1, T2, and T3 and compared model fit for solutions with one to five classes (Bengt Muthén, 2003; Nylund, Asparouhov, & Muthén, 2007). All analyses were conducted in Mplus version 8 (Muthén & Muthén, 2017) using a robust full information maximum likelihood (FIML) estimation procedure for handling missing data which assumes missing data are unrelated to the outcome variable (i.e., missing at random) (Enders, 2001). We considered multiple criteria to evaluate model fit, including the Bayesian Information Criterion (BIC), Lo–Mendell–Rubin adjusted Likelihood Ratio Test, and entropy values (B. Muthén, 2004; Bengt Muthén, 2003; Nylund et al., 2007). We also evaluated the substantive meaning of class solutions relative to theoretical accounts and prior findings (Bonanno, 2012; Bonanno, Westphal, & Mancini, 2011). We initially compared fit for unconditional models without covariates using conventional indices that penalize more complex models when they fail to provide a better fit to the data (Lo, 2001; Bengt Muthén, 2003; Nylund et al., 2007). However, consistent with recommendations, we then included covariates as predictors of class membership in conditional models. Covariates were age, sex, education, income, living alone, and living in the US. This step is recommended because non-significant relationships between covariates and class membership may indicate an incorrect local solution (Bengt Muthén, 2003).

We next compared demographic factors, COVID-19 stressors and COVID-19 worries between the anxiety trajectory classes. We first used univariate ANOVA to compare the derived trajectory groups on a variety of demographic factors (age, gender, race, income, education, living alone, and living in the US). We then used multinomial logistic regression models to test whether T1 COVID-19 stressors (e.g., testing positive, knowing someone who died) and worries (i.e., worries about contracting, financial burden of the pandemic) predicted class membership controlling for demographic variables. Finally, we used multinomial logistic regression models to test whether T1 resiliency (e.g., emotion regulation, harmony in close relationships) or risk (e.g., prior anxiety diagnosis of anxiety, sleep problems) factors predicted class membership, controlling for demographic factors. Note that as a sensitivity analysis, we repeated the LGMM analyses using just participants from the US.

## Results

### Identifying trajectories of anxiety

Descriptive statistics for study variables at T1, T2, and T3 assessments are presented in **Table 1**. Bivariate correlations between anxiety assessed at each time point and all study variables at T1 are presented in supplemental **Table S1**. The fit statistics and theoretical interpretability for the unconditional and conditional models showed that a four-class conditional model provided the best fit of the data (see supplemental **Table S2**). The majority of participants (*n* = 961; 75.08%) showed stable low anxiety over time and a non-significant slope (*resilient*) (*B* = .23, *SE* = .47, *p* = .62). The next largest group (*n* = 149; 11.64%) displayed stable high anxiety over time (*chronic*) (*B* = .83, *SE* = .49, *p* = .09). A smaller group (*n* = 96; 7.50%) began with high levels of anxiety that decreased over time (*recovered*) (*B* = −5.03, *SE* = 1.13, *p*<.05). Finally, a small group of participants (*n* = 74; 5.78%) began with lower levels of anxiety that increased over time (*delayed*) (*B* = 6.33, *SE* = 1.24, *p*<.01). **Figure 1** depicts the actual classes trajectories. See supplemental **Table S3** for details of the intercept and slope factors. When the sample was restricted to US participants, a similar four-class solution also emerged (see supplemental **Tables S4** and **S5**).

**Table 1.** Sample descriptive statistics (*n*=1362)

|  | % of Participants (n) |
| --- | --- |
| **Sex** |  |
| Female | 82.5% (n=1123) |
| Male | 17.5% (n=238) |
| Missing | 0.1% (n=1) |
| **Race** |  |
| White | 87.4% (n=1191) |
| Other | 10.6% (n=144) |
| Missing | 2.0% (n=27) |
| **Ethnicity** |  |
| Hispanic | 3.9% (n=53) |
| Non-Hispanic | 91.3% (n=1244) |
| Missing | 4.8% (n=65) |
| **Income** |  |
| < 100k | 46.9% (n=639) |
| ≥ \$100k | 47.4% (n=645) |
| Missing | 5.7% (n=78) |
| **Education** |  |
| Undergraduate Degree or Lower | 39.2% (n=534) |
| Master's Degree or Higher | 60.6% (n=826) |
| Missing | 0.1% (n=2) |
| **Living Arrangement** |  |
| Living Alone | 20.0% (n=272) |
| Living with Others | 80.0% (n=1089) |
| Missing | 0.1% (n=1) |
| **Country of Residence** |  |
| USA | 78.1% (n=1064) |
| Israel | 16.3% (n=222) |
| Other Countries | 5.6% (n=76) |
| Missing | 0% % (n=0) |
| **Past Diagnosis** |  |
| Anxiety | 24.5% (n=334) |
|  | **Mean (SD)** |
| **Age** |  |
| In years | 41.02 (13.67) |
| **GAD-7 Scores** |  |
| Time 1 | 6.26 (4.84) |
| Time 2 | 6.12 (5.03) |
| Time 3 | 5.96 (5.02) |

*Note.* Sample consists of participants who completed the GAD-7 for at least two time points. . GAD7 = Generalized Anxiety Disorder-7.

**Figure 1.**
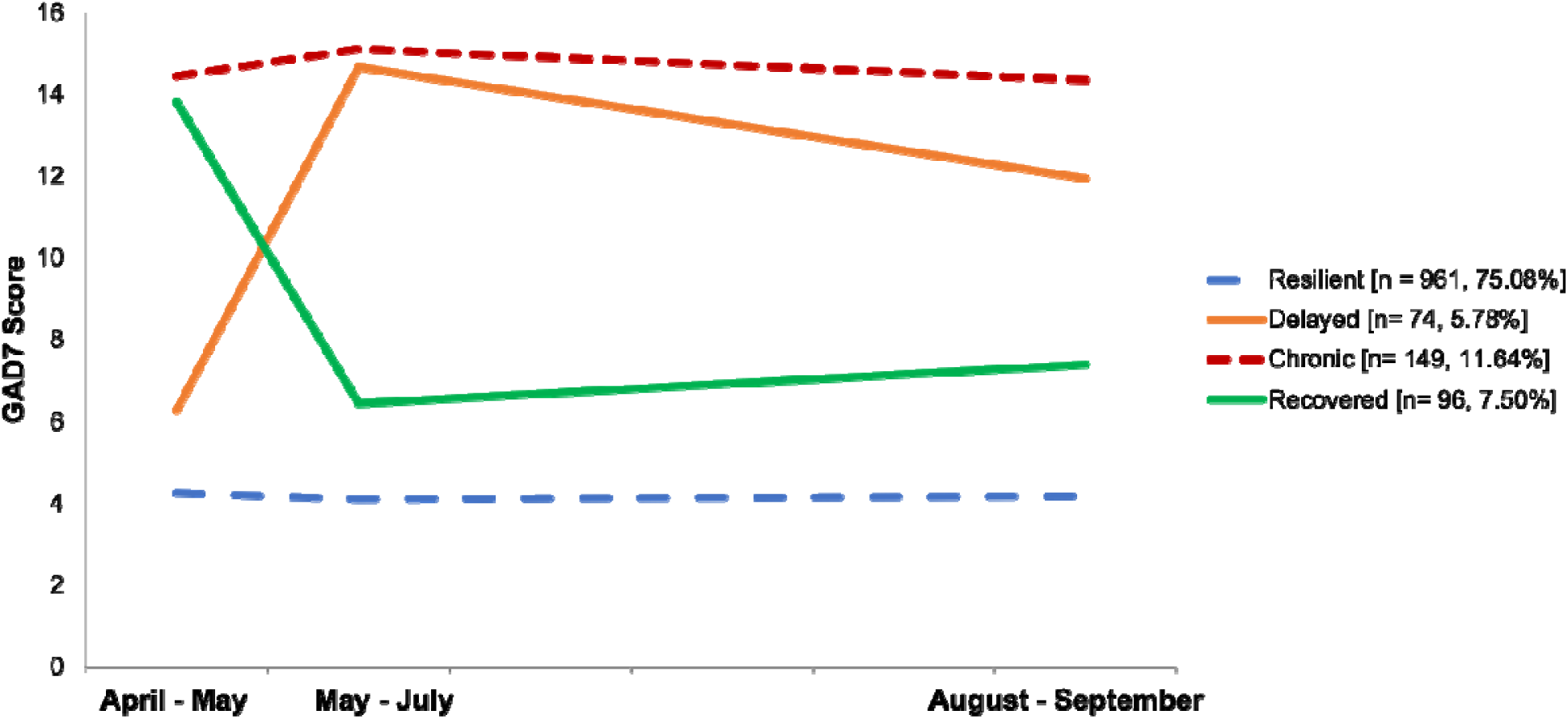
Mean Anxiety Scores Across the Study Period for the Four Anxiety Trajectory Groups *Note*. Anxiety was assessed using the GAD-7. Covariates were included on the intercept and slope factors and as predictors of class membership. Covariates included participant age, sex, education (0 = less than a Master’s degree; 1= Master’s degree or higher), income, living alone, and living in the US. All models were run in Mplus version 8. Model results remained unchanged when the sample was restricted to just the US participants.

### Characteristics of anxiety trajectories

#### Demographic characteristics

Resilient participants were more likely to be older, male, non-Hispanic, more educated, and less likely to live in the United States relative to the other trajectory groups (**Table 2**). For descriptive statistics of additional demographic variables see supplemental **Table S6**. Subsequent multinomial logistic regression and MANCOVA models accounted for the effects of demographic factors (i.e., age, sex, education, income, living alone, and living in the US) when testing putative differences between trajectory classes on COVID-19 stressors and worries and risk and resilience factors as measured at baseline assessment (T1).

**Table 2.**
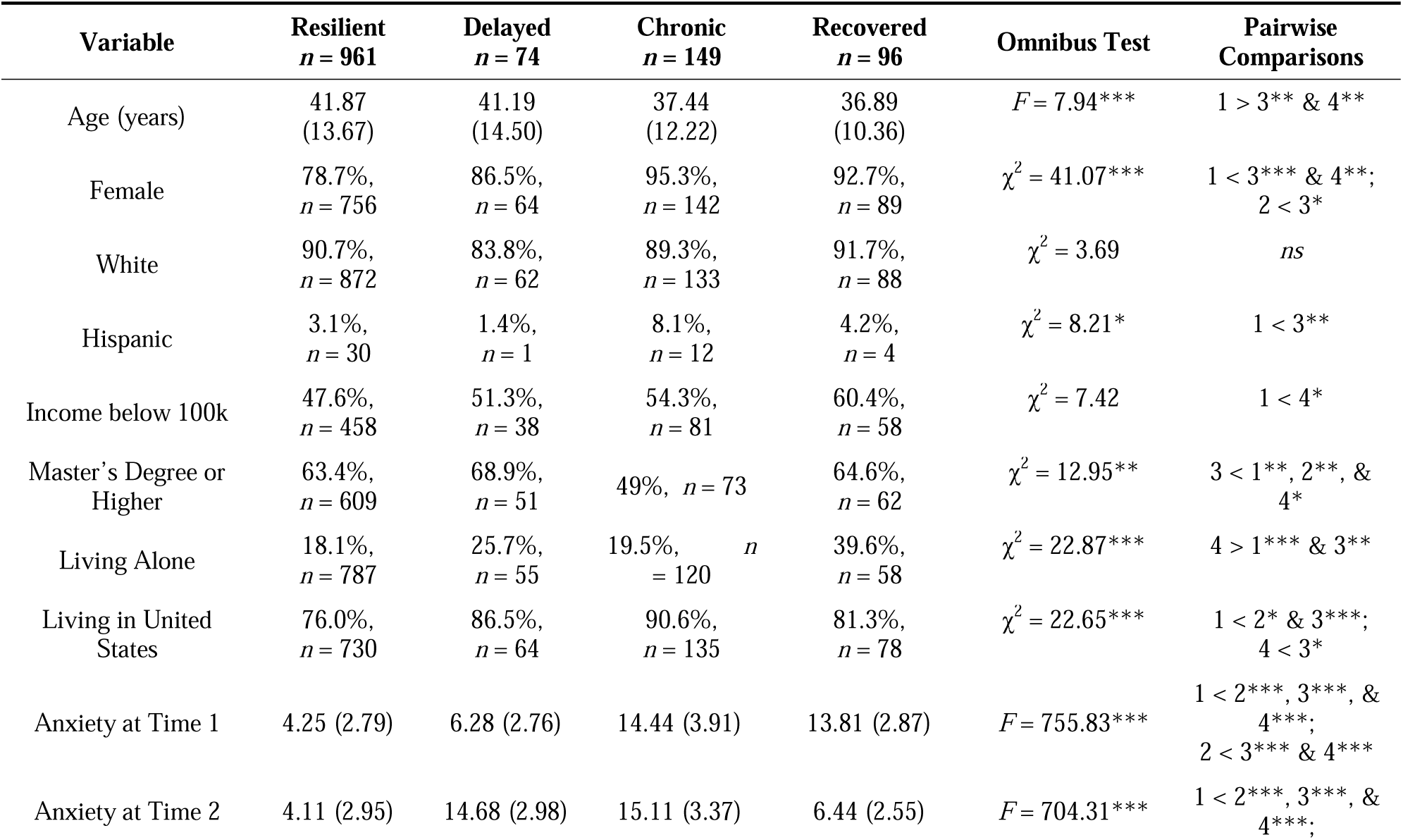
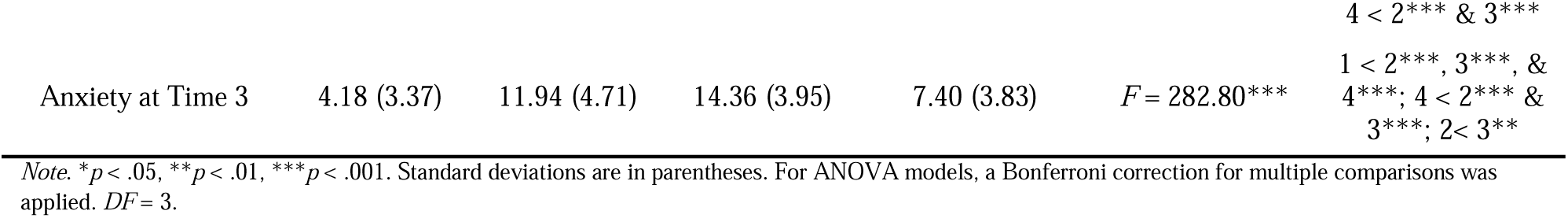
Descriptive Statistics and Comparisons of the Anxiety Trajectory Groups, Odd Ratios and Confidence Intervals [95% CI] for Multinominal Logistic Regression. Cumulative COVID stressors and Worries at T1 Differentiation among Trajectory Classes Membership

### Prospective predictors of anxiety trajectories

#### COVID-19 stressors

Multinomial logistic regression models that controlled for demographic factors revealed no significant differences between the trajectory classes on the basis of COVID-19 stressors at baseline. However, participants in both the chronic and recovered trajectories experienced significantly higher T1 COVID-19 worries about contracting/family contracting COVID-19 and the financial burden of the pandemic when compared to resilient participants. In addition, recovered participants experienced significantly higher worry about family contracting the virus and the financial burden of the pandemic relative to resilient participants. Finally, the chronic trajectory participants reported significantly higher worry about the financial burden of the pandemic compared to the recovered trajectory participants (**Table 3**). Similar findings emerged when comparing COVID-19 stressors and worries assessed at T2 and T3 (see supplemental **Tables S7 and S8**).

**Table 3.** Odd Ratios and Confidence Intervals [95% CI] for Multinomial Logistic Regressions Predicting Anxiety Trajectory from Cumulative COVID Exposures and Worries, Risk Factors and Individual Resiliency

| Predictor Variable | Resilient vs.<br>Delayed | Resilient vs.<br>Chronic | Resilient vs.<br>Recovered | Delayed vs.<br>Chronic | Delayed vs.<br>Recovered | Chronic vs.<br>Recovered |
| --- | --- | --- | --- | --- | --- | --- |
| Covid Exposures |  |  |  |  |  |  |
| Cumulative Exposures | 1.25 [.67-2.32] | 1.11 [.68-1.81] | 1.31 [.76-2.24] | .89 [.43-1.84] | 1.05 [.49-2.27] | 1.18 [.62-2.26] |
| Covid Worries |  |  |  |  |  |  |
| Self-contracting | 1.22 [.93-1.61] | **1.42\* [1.04-1.81]** | 1.37 [1.04-1.79] | 1.17 [.83-1.63] | 1.12 [.78-1.60] | .96 [.69-1.34] |
| Family contracting | 1.25 [.96-1.63] | **1.51\* [1.15-1.97]** | **1.57\*\* [1.21-2.04]** | 1.20 [.85-1.70] | 1.25 [.89-1.77] | 1.04 [.74-1.47] |
| Financial burden | **1.36\*\* [1.12-1.66]** | **1.59\*\*\* [1.35-1.88]** | **1.26\* [1.04-1.52]** | 1.17 [.93-1.48] | .92 [.72-1.19] | **.79\* [.63-.98]** |
| Risk Factors |  |  |  |  |  |  |
| Past Diagnosis of Anxiety | 1.6 [.89-2.88] | **2.08\* [1.30-3.33]** | .73 [.41-1.31] | 1.3 [.67-2.50] | **.46\*\* [.22-.96]** | **.35\*\*\* [.19-.67]** |
| Perceived Health | 1.13 [.83-1.54] | .92 [.72-1.18] | 1.04 [.76-1.43] | .81 [.57-1.16] | .92 [.61-1.39] | 1.13 [.80-1.61] |
| Sleep | **1.13\*\*\* [1.08-1.18]** | **1.20\*\*\* [1.14-1.25]** | **1.18\*\*\* [1.12-1.24]** | **1.06\* [1.00-1.11]** | 1.04 [.99-1.10] | .99 [.94-1.04] |
| Resiliency Factor |  |  |  |  |  |  |
| Emotion Regulation | **.91\* [.85-.99]** | **.85\*\*\* [.79-.90]** | **.85\*\*\* [.80-.90]** | .92 [.85-1.01] | .92 [.85-1.01] | 1.00 [.93-1.07] |
| Self-Reliance | 1.06 [.91-1.22] | .99 [.92-1.08] | 1.03 [.94-1.14] | .94 [.81-1.10] | .98 [.83-1.15] | 1.04 [.93-1.160] |
| Trust in Close Relationships | .98 [.89-1.07] | .94 [.87-1.01] | .97 [.89-1.05] | .96 [.86-1.07] | .99 [.88-1.11] | 1.03 [.94-1.13] |
| Low Conflict in Close Relationships | .95 [.86-1.05] | **.92\* [.85-.99]** | .97 [.89-1.05] | .97 [.86-1.09] | 1.02 [.90-1.16] | 1.06 [.96-1.16] |
| Neighborhood Environment | .96 [.90-1.04] | .96 [.91-1.02] | 1.07 [.996-1.14] | 1.00 [.92-1.09] | **1.11\* [1.01-1.21]** | **1.11\* [1.03-1.20]** |

*Note.* A total of three models were run with the resilient, delayed, or chronic group as the reference group respectively to allow for all possible group comparisons. The model controlled for the following covariates: participant age, sex, race (White= 1, Non-White= 0), Ethnicity (Hispanic=1, not Hispanic= 0), Educational Attainment, Income, and Whether the Participant is living alone and living in the US.
\* $p < .05$ , \*\* $p < .01$ , \*\*\* $p < .001$

#### Risk factors

Multinomial logistic regression models revealed that resilient trajectory participants reported significantly fewer sleep problems at baseline relative to all other trajectories. Likewise, the delayed anxiety participants reported fewer sleep problems than the chronic anxiety participants. Participants in the resilient trajectory were significantly less likely to report having a prior (pre-COCID-19) anxiety disorder diagnosis compared to those in the chronic trajectory. Similarly, the recovered participants were significantly less likely to have a prior anxiety disorder diagnosis compared to the delayed participants **(Table 3 and Figure 2)**

**Figure 2.**
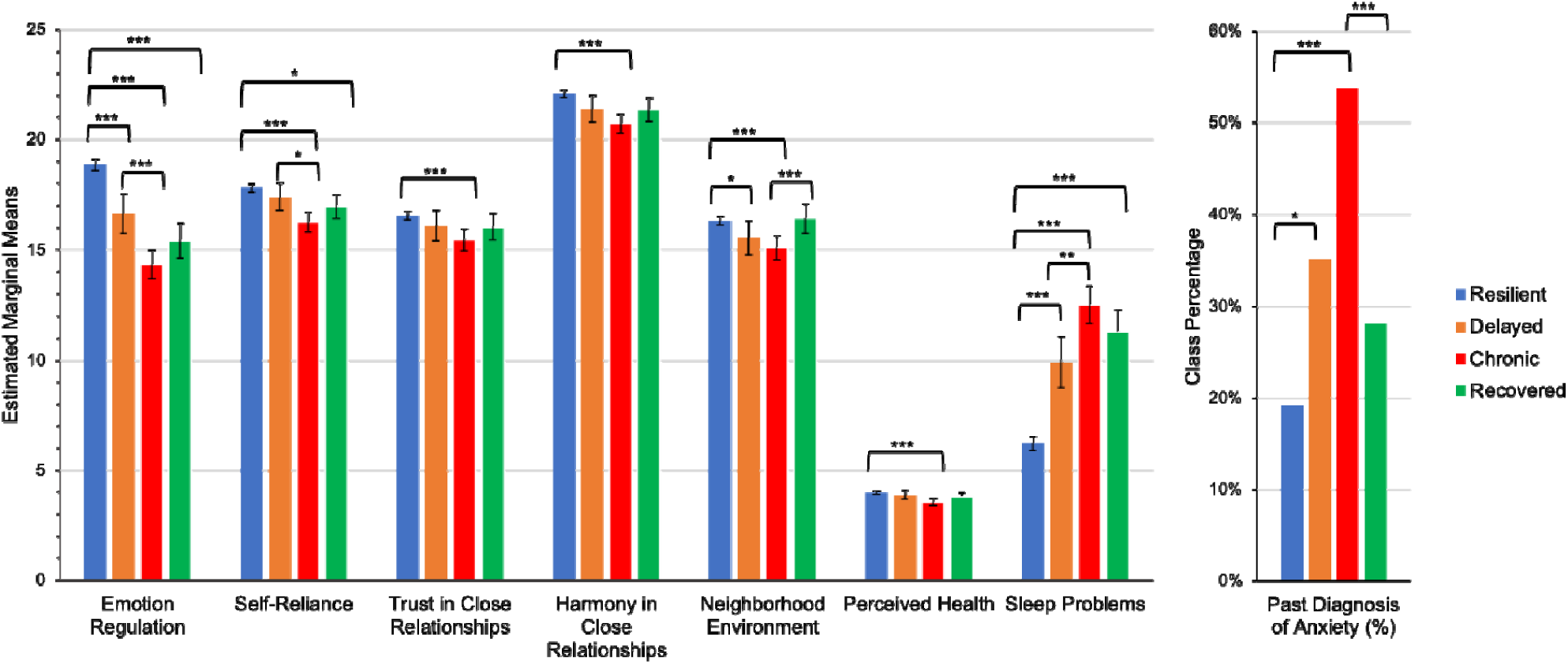
Estimated marginal mean scores and class percentages for the resilient, delayed, chronic, and recovered anxiety trajectory groups on measures of resiliency and risk at Time 1 *Note*. Error bars represent 95% CI. Significant effects were robust after applying a Bonferroni correction for multiple comparisons in SPSS vs. 26. Means were derived from a MANCOVA to aid interpretability. Results were similar when using the Multinomial Logistic Regressions and MANCOVA approach. See the supplement for results from the MANCOVA (Table S10). \**p* < .05, \*\**p* < .01, \*\*\**p* < .001.

#### Resiliency factors

Multinomial logistic regression models that controlled for demographic factors revealed that the resilient trajectory participants showed significantly greater emotion regulation relative to all other trajectories. In addition, resilient participants reported significantly lower hostility in close relationships relative to the chronic trajectory. Recovered participants reported significantly higher levels of positive neighborhood environment relative to delayed and chronic trajectories participants **(Table 3 and Figure 2)**.

## Discussion

We identified distinct anxiety trajectories across the first six months of the COVID-19 pandemic. The patterns that emerged were consistent with theoretical expectations, revealing four distinct patterns of response. The most prevalent response in our study cohort was a resilient trajectory of low stable anxiety, suggesting that most individuals evidenced modest distress reactions, even in the face of the highly disruptive events of the COVID-19 pandemic. However, other distinct responses patterns also emerged, including a pattern of recovery from initially high levels of anxiety, delayed or worsening anxiety, and chronically elevated anxiety. Importantly, although the course of the pandemic changed markedly over time, as cases, deaths, lockdowns, and mask-wearing mandates varied over the study period, the vast majority of participants exhibited little change in anxiety levels, with more than 86% of the sample remaining stable in their anxiety levels (i.e., in the resilient or chronic trajectories). Our findings suggest the robust capacity for a large majority of individuals to remain resilient (i.e., not develop significant anxiety symptoms), even in response to an acute and uncertain stressor. However, a subset of individuals appeared vulnerable to persistent and worsening anxiety over time. We also identified several key demographic and risk and resilience factors that differentiated group membership, highlighting potential avenues for future personalized interventions.

From a population perspective over time, one can view the emergence of the COVID-19 outbreak as having potentially profound negative effects on mental health. However, prior research on acute and chronic stress exposures suggest that effects will show considerable variation across individuals, time, and contexts (Mancini, 2020). One way to assess this variation is to examine anxiety as an index of stress-reaction (Adhikari et al., 2015) and resilience as a stable outcome and positive adaptation to adversity (Bonanno, 2004). We used latent group-based trajectory modeling to identify subpopulations of individuals who show different patterns over time on a repeated measures outcome and the characteristics associated with resilient outcomes (Muthen, 2004). We found that a considerable majority of participants (75%) were resilient, with stable low levels of anxiety over time. Our findings are consistent with prior studies that have documented resilience as the most common pattern in response to a wide variety of stressors, including school shootings (Mancini, Littleton, & Grills, 2016), combat (Maguen et al., 2020), traumatic injury (deRoon-Cassini, Mancini, Rusch, & Bonanno, 2010) and bereavement (Mancini et al., 2011). The findings are also consistent with a longitudinal cohort study of 1296 participants in Australia in which similar anxiety trajectory patterns emerged, including a resilient pattern characterizing 77% of the sample. In addition, a recent review and meta-analysis of longitudinal studies on the psychological impact of COVID-19 pandemic lockdowns suggested that even in the face of recent lockdowns, a major stress that implicated people’s life, most individuals retain their capacity for psychological adaptation (Prati & Mancini, 2021).

Importantly, we were able to document differences between participants in the different trajectory classes on the basis of demographic factors, worries about and stressors associated with COVID-19, and baseline risk and resilience characteristics. In line with studies from current pandemic (Filgueiras & Stults-Kolehmainen, 2021; Sherman, Williams, Amick, Hudson, & Messias, 2020) and as documented in previous research on resiliency, (Bonanno, Romero, & Klein, 2015), a number of contextual variables were significantly associated with better outcomes; including older age, male gender, and greater level of education. Indeed, men generally report lower levels of psychological distress (Löwe et al., 2008), yet another possible explanation could be the increased burden around childcare during school closures, which was especially high among working mothers (Power, 2020). Findings regarding older adults may reflect their tendency to develop strengths through a lifetime of experiences, or their ability to negotiate through challenges better than younger adults (Charles, 2010).

In terms of stressors to and worries about the pandemic, we found no differences between groups on the basis of COVID-19 baseline stressors. That is, groups did not differ in their exposure to the virus or the participant’s occupational consequences. In terms of worries, we found that participants in the chronic anxiety trajectory reported significantly more worries related to COVID-19, notably in relation to financial burdens, as well as concerns for self and others contracting the virus, and suffered from poorer sleep quality, replicating prior findings in study of German participants (Gilan et al. 2020). Sleep represents a neurobiological balance between arousal and de-arousal and considered as a basic dimension for brain function and mental health (Harvey, Murray, Chandler, & Soehner, 2011). Sleep quality was previously found to mediate the association between COVID-related stressors and mental health outcomes in a large community samples of adults from the United States and Israel during the early stages of the pandemic, highlighting the central role that sleep plays in promoting resilience in the face of stress during COVID 19 pandemic (Coiro et al., 2021).

Finally, we found that more than 50% of participants in the chronic anxiety group had reported a past diagnosis of anxiety. This finding is consistent with prior studies suggesting that the negative effects of the COVID-19 pandemic appear more pronounced among vulnerable populations (Pan et al., 2021). Thus, it is essential to implement programs that can overcome systemic barriers of care and decrease global health disparities in vulnerable populations.

In terms of resilience, we found that emotion regulation was significantly higher among participants in the resilient trajectory compared to all other trajectories. Notably, emotion regulation scores significantly differed between participants in the resilient trajectory and those in the delayed anxiety trajectory, suggesting that this capacity may be important to sustain low anxiety levels throughout longitudinal exposure to the pandemic related stress (i.e., 6-months of study period). Broadly defined, emotion regulation includes the ability to identify and accept emotional experiences, control impulsive behaviors when distressed, and flexibly modulate emotional responses as situationally appropriate (Renna, Quintero, Fresco, & Mennin, 2017). Low levels of emotion regulation have previously been linked to risk for anxiety and depression (Chen and Bonanno 2021), as well as externalizing problems (Cappadocia, Desrocher, Pepler, & Schroeder, 2009; Mitchell, Robertson, Anastopolous, Nelson-Gray, & Kollins, 2012). Hence, it is not merely a signal for anxiety, but represent instead, a trans-diagnostic marker for psychopathology (Aldao, Nolen-Hoeksema, & Schweizer, 2010; Beauchaine & Thayer, 2015). High emotion regulation capacity, biologically reflected by prefrontal inhibition, is considered as a key mechanism in psychological health, hence crucial for adaptive functioning (Aldao et al., 2010; Gross, 2007). Our findings support pre-pandemic data suggesting that emotion regulation serves as mediator of the relationship between resilience and distress (Vaughan et al., 2019), and expand latest findings suggesting that individual differences in emotion regulation prospectively predict early COVID-19 related acute stress (Tyra, Griffin, Fergus, & Ginty, 2021). Specifically, we suggest that interventions aimed at emotion regulation might be especially warranted as a modality to enhance resilience (Lee et al., 2020; Renna, Fresco, & Mennin, 2020).

A second resilience factor that differentiated between the chronic and resilient groups was hostility in close relationships, with significant low hostility features reported by the resilient trajectory participants. This findings might be indicative of the different interpersonal challenges facing individuals during the pandemic, with people spending many weeks under lockdown with close others, for better or worse (Gadermann et al., 2021). That is, close relationships coping competences can be a way to enhance ability to deal with the stress, concerns and adverse events related to the pandemic (Prime, Wade, & Browne, 2020). In particular, dyadic coping had been found to play a critical role in stress reduction and in restoring psychological well-being during the COVID-19 emergency (Donato et al., 2021). Finally, we found that a more positive neighborhood environment, a resilience factor reflecting feelings of personal safety and community cohesion (Mujahid, Diez Roux, Morenoff, & Raghunathan, 2007), was significantly higher among recovered participants compared to participants in the delayed and chronic anxiety trajectories, highlighting the importance of social cohesion and community support in the face of the pandemic (Fone et al., 2014). Indeed resilience may be supported by the enhanced availability of social support after shared stressors (Mancini, Westphal, & Griffin, 2021). Relevant to both individual and community aspects of resiliency, recent research suggests that community resilience, in addition to individual levels resilience, informs resilient outcomes and capacity to cope with threats (Kimhi, Marciano, Eshel, & Adini, 2020; Ungar & Theron, 2020).

A strength of the current study is the leveraging of a large sample with prospective longitudinal data collected over three time points at a critical period of the pandemic. However, our findings should be considered alongside several important study limitations. First, we used an on-line “snow-ball” recruitment method, which reduced the representativeness of our sample and the generalizability of the findings. Specifically, people who complete online surveys differ from people who do not, and thus the proportions of people in each trajectory should not be taken as population-level estimates (Pierce, McManus, et al., 2020). Second, our cohort included participants from the US and Israel, countries characterized by considerable differences in virus spread, government restrictions, and health care. Indeed, living in Israel predicted membership in the resilient group. Notably, however, at time T1 data collection, the restrictions on residents were similar in both Israel and the US. Moreover, a recent study in the US, the United Kingdom, and Israel similarly found that Israeli participants exhibited lower levels of general anxiety compared to others (Bareket-Bojmel, Shahar, & Margalit, 2020), which might reflect cultural differences in expressing anxiety symptoms. Importantly, the trajectory patterns were largely robust to these differences, and similar findings emerged when the sample was restricted to only the US participants.

In sum, we found distinct patterns of adaptation to the COVID-19 pandemic in our longitudinal cohort. These patterns were prospectively predicted by dispositional resilience factors and by COVID-19 stressors, suggesting that self-reported capacities for resilience and resources for resilience predict longitudinal outcomes. Together, these findings suggest most people show a robust capacity for resilience to the pandemic, but they also suggest that COVID-19 stressors contributed to worse adjustment among a subset of vulnerable individuals. Intriguingly, given that dispositional resilience factors prospectively predicted resilient outcomes, our findings highlight future targets for intervention that can help to reduce the mental health burden following major global stressors.

## Data Availability

Data collected for this study includes individual participants data. Data cannot be publicly
accessible due to Institutional Review Board guidelines. We are open to collaborations with
other researchers upon contacting us.

## Acknowledgements

We thank participants of covid19resilience.org for their contribution to data generation.

## Ethical standards

The authors assert that all procedures contributing to this work comply with the ethical standards of the relevant national and institutional committees on human experimentation and with the Helsinki Declaration of 1975, as revised in 2008. The study was approved by the Institutional Review Board of the University of Pennsylvania.

